# Radical Vulvectomy with Rectus Abdominis Flap Reconstruction for Recurrent Vulvar Cancer Post-Radiotherapy: Clinical Efficacy and Functional Outcomes

**DOI:** 10.64898/2026.05.04.26352038

**Authors:** Guixiang Zhang, Chongqing Gao, Tao Wang, Zhen Zhang, Zhi Zhang, Zikang Wang, Shuo Li, Xinyao Zhang, Gangcheng Wang

**Author notes:** **Corresponding author.** Contact Information: Dr. Gangcheng Wang, Department of Abdominal and Pelvic Tumor Surgery, The First Affiliated Hospital of Zhengzhou University. Guixiang Zhang, Chongqing Gao, Tao Wang, and Zhen Zhang contributed equally to this work and share first authorship. **all author details and affiliations** Gangcheng Wang, The First Affiliated Hospital of Zhengzhou University, Department of Abdominal and Pelvic Tumor Surgery, Zhengzhou, Henan Province, China. 450052. Guixiang Zhang, The First Affiliated Hospital of Zhengzhou University, Department of Abdominal and Pelvic Tumor Surgery, Zhengzhou, Henan, China. 450052. Chongqing Gao, The First Affiliated Hospital of Zhengzhou University, Department of Abdominal and Pelvic Tumor Surgery, Zhengzhou, Henan, China. 450052. Tao Wang, The First Affiliated Hospital of Zhengzhou University, Department of Abdominal and Pelvic Tumor Surgery, Zhengzhou, Henan, China. 450052 Zhen Zhang, The First Affiliated Hospital of Zhengzhou University, Department of Abdominal and Pelvic Tumor Surgery Zhengzhou, Henan, China.450052. Zhi Zhang, The First Affiliated Hospital of Zhengzhou University, Department of Abdominal and Pelvic Tumor Surgery, Zhengzhou, Henan, China. 450052 Zikang Wang, The First Affiliated Hospital of Zhengzhou University, Department of Abdominal and Pelvic Tumor Surgery, Zhengzhou, Henan, China. 450052. Shuo Li, The First Affiliated Hospital of Zhengzhou University, Department of Abdominal and Pelvic Tumor Surgery, Zhengzhou, Henan, China. 450052. Xinyao Zhang, The First Affiliated Hospital of Zhengzhou University, Department of Abdominal and Pelvic Tumor Surgery, Zhengzhou, Henan, China. 450052. **Guarantor of the article:** Dr. Gangcheng Wang.

## Abstract

**Objective:** Recurrent vulvar cancer after radiotherapy is a devastating condition that severely impairs patients’ quality of life. Although radical vulvectomy can improve survival and quality of life in primary cases, patients with post-radiation recurrence are at high risk for severe wound-related complications. To address this challenge, our team adopted a combined approach of radical vulvectomy and rectus abdominis flap reconstruction, which has yielded favorable therapeutic outcomes.

**Methods:** This retrospective study enrolled patients with radiotherapy-recurrent vulvar cancer who underwent radical vulvectomy combined with rectus abdominis flap reconstruction between 2023 and 2024. The primary endpoint was the change in quality of life, assessed using the EORTC QLQ-C30 and QLQ-VU34 questionnaires.

**Results:** Eleven patients underwent radical vulvectomy with rectus abdominis flap reconstruction. The median patient age was 67 years, and the median BMI was 28.98. The median operative time and intraoperative blood loss were 210 minutes and 400 mL, respectively. All patients achieved R0 resection. Nine patients experienced only minor complications (Clavien - Dindo grade II), while two patients developed complications requiring intervention under local anesthesia (grade IIIa). No wound infections, wound dehiscence, or donor-site complications were observed. Postoperatively, a significant improvement in quality of life was evident across all patients, particularly in the domains of physical functioning, pain reduction, and overall global health status (all P < 0.05).

**Conclusions:** Radical vulvectomy combined with rectus abdominis flap reconstruction significantly improves quality of life and oncological outcomes in patients with radiotherapy-recurrent vulvar cancer. This combined approach represents a valuable and safe treatment option that merits broader clinical implementation, preferably within a multidisciplinary team setting.

## Introduction

Vulvar cancer, a relatively rare malignancy of the female external genitalia, accounts for 5–8% of gynecological cancers.^1,2^ Recent epidemiological data indicate a concerning upward trend in its incidence, particularly among women under 60 years of age.^3,4^ Its pathogenesis is multifactorial, involving well-documented risk factors such as human papillomavirus (HPV) infection, vulvar intraepithelial neoplasia (VIN), chronic inflammatory conditions, immunosuppression, genetic susceptibility, advanced age, tobacco use, racial disparities, and occupational hazards. Notably, high-risk HPV subtypes—especially HPV16 and HPV18—have been strongly linked to vulvar carcinogenesis.^5,6^

The histopathological classification of vulvar cancer encompasses several subtypes, including squamous cell carcinoma (SCC), malignant melanoma, basal cell carcinoma, adenocarcinoma, Paget’s disease of the vulva, verrucous carcinoma, and sarcoma. Among these, SCC is the predominant variant, accounting for over 95% of cases.^7^ The clinical manifestations of vulvar cancer are diverse; common symptoms include persistent vulvar itching or pain, lumps or nodules, ulcers, bleeding or increased discharge, worsening pain during urination and sexual intercourse, and lower limb swelling or dysfunction.^8^ Because of its anatomically sensitive location and prominent symptoms, vulvar cancer causes substantial psychological distress and markedly impairs patients’ quality of life.

Primary vulvar cancer is mainly treated with surgical excision or radiation therapy, both of which are effective for early-stage disease. However, studies show that 12% to 37% of patients experience recurrence within two years of initial treatment.^9^ In particular, the recurrence rate may be as high as 40% to 50% in patients treated with surgery alone.^10^ For patients with recurrent vulvar cancer who have not previously received radiation therapy, options such as radiation therapy or reoperation may be considered. However, the National Comprehensive Cancer Network (NCCN) has not provided detailed treatment recommendations for patients whose cancer recurs after radiation therapy; in the 2024 NCCN guidelines, radical vulvectomy is listed as an acceptable option.^1^ Notably, prior irradiation compromises cutaneous regeneration, increasing the risk of postoperative complications—such as chronic wound dehiscence or delayed healing—in patients undergoing radical resection.^11^ To address the potential complications associated with radical vulvectomy in patients with recurrent vulvar cancer after radiotherapy, our research team explored the feasibility of combining radical vulvectomy with flap functional reconstruction. Among various flap reconstruction techniques—such as rectus abdominis muscle flap, gluteus maximus muscle flap, gracilis muscle flap, and anterolateral thigh flap—the rectus abdominis muscle flap is the preferred choice. This is due to its advantages of rich blood supply, sufficient tissue volume, good pliability, lower surgical technical demands, and fewer donor-site complications.^12,13^ Based on these considerations, our team proposed a treatment strategy combining radical vulvectomy and functional reconstruction using the rectus abdominis muscle flap for patients with recurrent vulvar cancer after radiotherapy. Currently, few studies have explored this treatment modality. We therefore conducted a clinical study involving 11 patients who underwent this combined approach. This study aims to evaluate its clinical efficacy, with a particular focus on postoperative complications and quality of life outcomes, thereby providing valuable insights for clinical practice in this field.

## Materials and Methods

### Study design

The objective of this study was to retrospectively evaluate the clinical outcomes of radical vulvectomy combined with rectus abdominis flap functional reconstruction in patients with recurrent vulvar cancer after radiotherapy. A total of 11 patients between 2023 and 2025 were included. All surgical procedures were performed and documented by our team. Inclusion criteria were limited to patients with radiotherapy-recurrent vulvar cancer who underwent radical vulvectomy and rectus abdominis flap reconstruction. Patients with primary disease, distant metastases, no prior radiotherapy, or those who received other reconstructive approaches were excluded. The study was approved by the institutional ethics committee of the leading participating hospital (Approval No. SB-2024-204-001). The study used retrospectively collected, anonymized data, and written informed consent was obtained from all patients. All data processing complied with relevant institutional data security and privacy protocols.

In this study, patients’ quality of life was assessed using the EORTC QLQ-C30 (version 3.0) and the vulvar cancer-specific module QLQ-VU34.^14,15^ The QLQ-C30 consists of 30 items covering five functional scales (physical, role, cognitive, emotional, and social functioning), three symptom scales (fatigue, pain, nausea and vomiting), a global health status/QoL scale, and six single items. Higher scores on the functional and global health scales indicate better functioning and quality of life, whereas higher scores on symptom scales reflect greater symptom burden. The QLQ-VU34 is a supplementary module designed to assess disease- and treatment-related symptoms specific to vulvar cancer. It comprises ten multi-item scales evaluating how the clinical manifestations of vulvar cancer affect patients’ quality of life, and is scored using the same standardized method as the QLQ-C30.

### Data collection

This study was reported in accordance with the STROBE guidelines for observational studies to ensure methodological quality and transparency.^16^. All clinical data were retrospectively retrieved from the electronic medical record systems of the participating centers, with data anonymized to protect patient privacy. The data were obtained through a detailed review of hospital admission records, follow-up records, and imaging reports. Collected variables included patient demographics, recurrence status, prior history of radiotherapy or chemoradiotherapy, clinical presentation (including ulcer size, pain severity, and other notable symptoms), surgical details (including specifics of the flap reconstruction), and postoperative complications and survival.

### Definitions

Radical vulvectomy is a radical surgical procedure for patients with vulvar cancer.^17,18^ The primary goal is to excise the primary tumor with an adequate margin of surrounding tissue, which may include the clitoris, vaginal wall, urethra, or anorectum depending on tumor extent. Excision typically extends to the deep thigh fascia, pubic symphysis, and deep perineal musculature to ensure complete tumor removal. Inguinal lymph node assessment is performed through a separate incision when indicated. The surgical approach should be tailored to the tumor’ s location, size, and histological characteristics, balancing radical resection with preservation of physiological function and appearance to optimize postoperative quality of life.

Flap reconstruction is a surgical technique that involves the transfer of skin and associated subcutaneous tissue, primarily used for repairing defects caused by disease or trauma. Flaps are categorized into pedicled flaps, which retain their vascular pedicle and are tunneled to the recipient site, and free flaps, which are completely detached and require microsurgical anastomosis of blood vessels to the recipient site. Commonly used flaps for perineal reconstruction include the pedicled rectus abdominis muscle flap, gluteus maximus muscle flap, gracilis muscle flap, and anterolateral thigh flap. The rectus abdominis flap is often preferred due to its robust blood supply, substantial tissue volume, and favorable pliability, which facilitate repair of large perineal defects.

### Endpoints

The primary endpoint was the change in quality of life, as assessed by the EORTC QLQ-C30 (version 3.0) and QLQ-VU34 questionnaires, from baseline to the most recent follow-up. Secondary endpoints included the rate of postoperative complications (classified according to the Clavien-Dindo system, with grade ≥3 defined as major complications), length of hospital stay, overall survival, and disease recurrence.^19^

### Statistical analysis

Standardized scores for the five functional scales, three symptom scales, six single-item measures, and the global health status/QoL scale of the EORTC QLQ-C30, as well as the ten multi-item scales of the QLQ-VU34, were calculated according to the EORTC scoring manual. Preoperative and postoperative scores were compared using the paired-samples t-test for normally distributed variables and the Wilcoxon signed-rank test for non-normally distributed variables. A two-sided P < 0.05 was considered statistically significant. All analyses were performed using SPSS 27.0.

## Results

### Patient Characteristics and Perioperative Data

Between April 2023 and December 2024, a total of 11 patients with radiotherapy-recurrent vulvar cancer underwent radical vulvectomy with rectus abdominis flap reconstruction at our participating centers. All patients had previously received radiotherapy as part of their initial treatment, and 10 had also received chemotherapy. The median patient age was 67 years (range: 50–76 years), and the median BMI was 28.98 (classified as overweight per WHO criteria). Following radical vulvectomy, the median skin defect area was 82.5 cm^2^. The median operative time was 210 minutes (range: 180–300 minutes), and the median intraoperative blood loss was 400 mL. All patients achieved margin-negative (R0) resection, and postoperative pathology confirmed squamous cell carcinoma in all cases. Detailed clinical and surgical characteristics are summarized in Table 1.

**Table 1.**
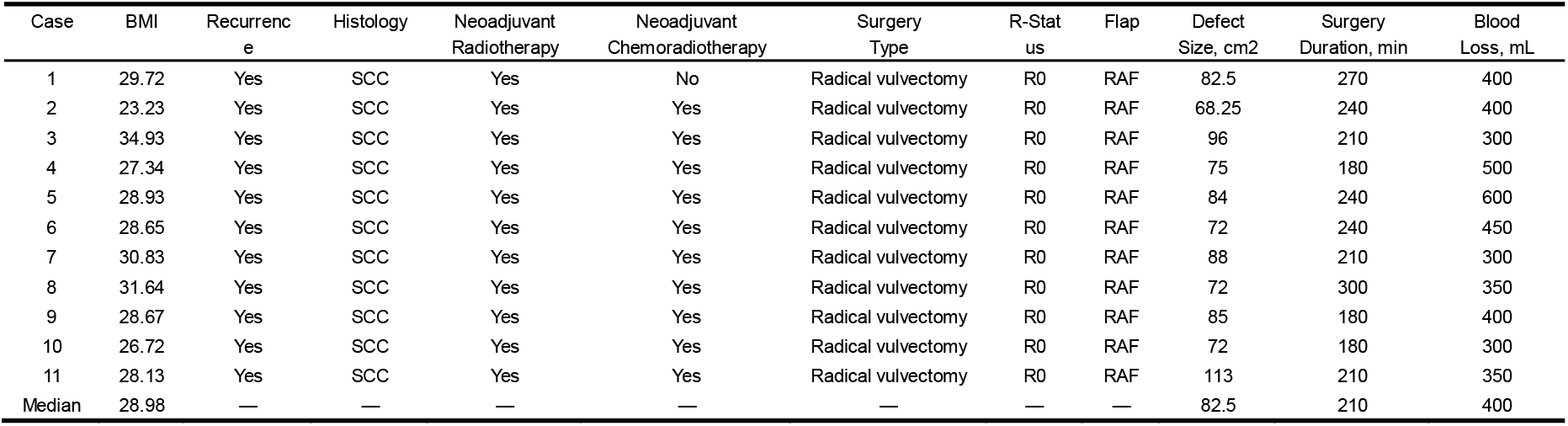
Clinical and surgical characteristics of the patients.

### Surgical methods

#### Radical vulvectomy

Patients were placed in the lithotomy position, and routine skin antisepsis and draping were performed. The surgical incision was designed 2–3 cm from the tumor edge. The perineal skin and subcutaneous tissues were incised following the tumor contour, followed by sequential dissection of the superficial perineal fascia and muscles.

Intraoperatively, the relationship between the tumor and the anterior rectal wall was assessed. If no invasion was present, adhesions were carefully separated using blunt and sharp dissection. If tumor invasion of the rectal wall was identified, the involved segment was resected to achieve R0 resection, and a prophylactic colostomy was performed. In cases of extensive rectal involvement, an abdominoperineal resection (APR) was performed.

The relationship between the tumor and the pubic bone was similarly evaluated. If the pubic bone was not involved, the anterior tumor margin was separated along the retropubic space, with careful dissection of the urethra while preserving the urethral sphincter to maintain urinary continence. If pubic bone invasion was present, the involved portion of the pubic bone or symphysis pubis was resected, and inguinal lymphadenectomy was performed.

The lateral tumor attachments were divided, and the tumor was mobilized along the medial aspect of the ischial tuberosity and the ischiopubic ramus. The pelvic floor muscles were dissected and resected according to tumor depth, allowing complete exposure of the lateral vaginal wall. The anterior vaginal wall was divided, the paravaginal tissues were fully exposed, and the posterior vaginal wall was separated from the rectal space. The tumor was completely excised, and the posterior vaginal wall was transected at the level of the cervical os.

The surgical field was thoroughly irrigated, and meticulous hemostasis was achieved. A catheter was placed through the urethral meatus to drain urine and to stent the neo-urethra. Representative images of the radical vulvectomy procedure are shown in Figure 1.

**Figure 1.**
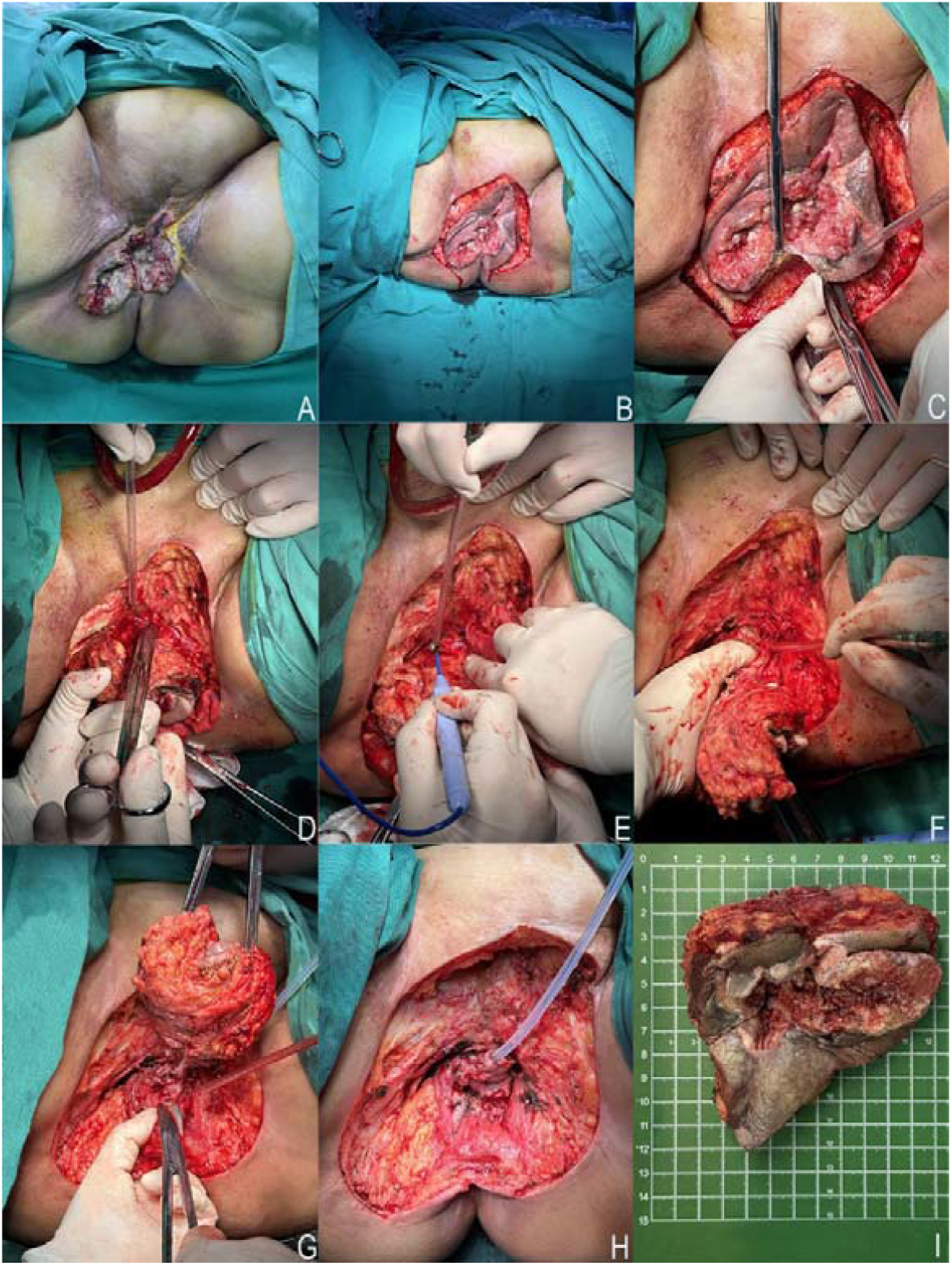
Representative images of a radical vulvectomy. (A) Representative example image of vulvar cancer. (B-G) Representative images of radical vulvectomy surgery. (H) Reconstruction of the artificial urethra. (I) Specimens that were surgically removed.

#### Rectus Abdominis Muscle Flap Reconstruction

The perineal skin defect was precisely measured, and an appropriately sized rectus abdominis myocutaneous flap was designed. The skin, subcutaneous tissue, and anterior rectus sheath were sequentially incised along the premarked flap margins. To prevent shearing, the anterior rectus sheath and subcutaneous tissue were intermittently sutured together, while the integrity of the posterior rectus sheath was carefully preserved.

Next, the attachments of the rectus abdominis muscle to the costal cartilages and xiphoid process were divided. The superior epigastric vascular pedicle—a terminal branch of the internal thoracic artery —was identified and ligated. The rectus abdominis muscle was then carefully elevated off the posterior rectus sheath in a craniocaudal direction, with meticulous preservation of the deep inferior epigastric artery and its perforating branches. The dissection continued inferiorly to completely release the pubic attachment of the muscle.

A subcutaneous tunnel was created anterior to the pubic symphysis, connecting the abdominal donor site to the perineal defect. The rectus abdominis flap was carefully transferred through this tunnel to the perineum, with vigilant protection of its vascular pedicle. The urethral orifice was reconstructed, and the flap was sutured to the perineal skin with interrupted sutures.

A synthetic mesh was placed to reinforce the abdominal donor site, and the abdominal wound was closed in layers. Representative images of the rectus abdominis flap reconstruction are shown in Figure 2.

**Figure 2.**
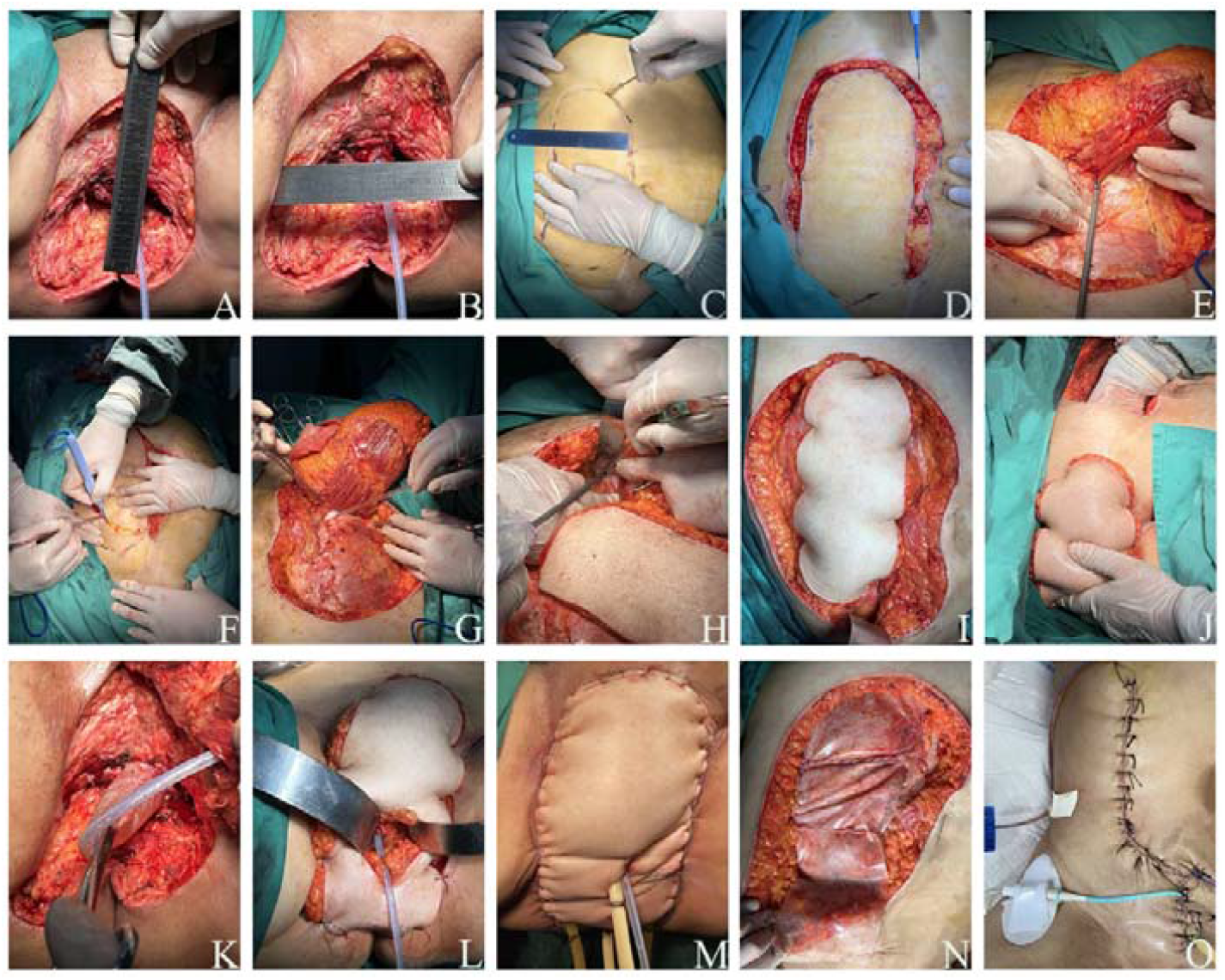
Representative images of a rectus abdominis muscle flap reconstruction. (A-C) Designing the rectus abdominis muscle flap. (D-J) Representative Images of the Reconstructive Surgery of the Rectus Abdominis Flap. (K-L) Reconstruction of the urethra. (M) Perineal trauma following repair of the rectus abdominis flap. (N-O) Repairing the donor site of rectus abdominis muscle flaps.

#### Complications

Postoperative complications were classified according to the Clavien–Dindo system. Nine patients (82%) experienced minor complications (grade II), primarily related to routine postoperative management (e.g., need for parenteral analgesics or nutritional support due to fasting). Two patients (18%) developed major complications (grade ≥3), both of which were grade IIIa and required reoperation under local anesthesia. No wound infections, wound dehiscence, or abdominal donor-site complications (e.g., incisional hernia, infection, dehiscence) were observed. The median length of hospital stay was 24 days.

The median follow-up duration was 13 months. At the last follow-up, all patients were alive, and the rectus abdominis flap had healed well in all cases. Ten patients remained disease-free, and one patient developed a pelvic recurrence. The complication profiles and surgical outcomes of all 11 patients are summarized in Table 2.

**Table 2.** Complications and outcome after Radical vulvectomy and rectus abdominis flap reconstruction.

| Case | Wound infection | Wound bleeding | Reoperation | Complication According to the Clavien-Dindo Classification | Hospital Stay, day | Long-Term Outcome | Duration of Follow-Up, month | Donor site complications |
| --- | --- | --- | --- | --- | --- | --- | --- | --- |
| 1 | No | No | No | II | 40 | Disease free | 3 | No |
| 2 | No | No | No | II | 35 | Disease free | 4 | No |
| 3 | No | No | Yes | IIIa | 27 | Pelvic recurrence | 19 | No |
| 4 | No | No | No | II | 24 | Disease free | 16 | No |
| 5 | No | No | No | II | 20 | Disease free | 9 | No |
| 6 | No | No | No | II | 21 | Disease free | 9 | No |
| 7 | No | No | No | II | 22 | Disease free | 18 | No |
| 8 | No | No | No | II | 26 | Disease free | 17 | No |
| 9 | No | No | No | II | 30 | Disease free | 8 | No |
| 10 | No | No | Yes | IIIa | 23 | Disease free | 13 | No |
| 11 | No | No | No | II | 20 | Disease free | 12 | No |
| Median | — | — | — | — | 24 | — | 13 | — |

#### Quality of life assessments

Quality of life was assessed pre- and postoperatively using the EORTC QLQ-C30 and QLQ-VU34 questionnaires. Compared with baseline, postoperative scores showed significant improvements across multiple QoL dimensions. Functional and global health scales. Patients’ mean scores for physical, role, emotional, cognitive, and social functioning, as well as global health status/QoL, were significantly increased (all P < 0.05), indicating marked improvement in functional status and overall well-being. Symptom scales. Significant reductions were observed in fatigue, pain, nausea and vomiting, dyspnea, insomnia, appetite loss, and constipation (all P < 0.05), reflecting effective symptom relief following surgery. No changes were observed in financial difficulty or diarrhoea scores after surgery. Vulvar cancer-specific symptoms. Patients reported significant decreases in vulvar skin changes, vulvar scarring, vulvovaginal discharge, vulvar swelling, and inguinal and leg lymphedema (all P < 0.05), underscoring the efficacy of the procedure in addressing disease-specific morbidity. Body image also improved significantly (P < 0.05), consistent with positive psychological adaptation. Detailed pre- and postoperative QoL scores for all 11 patients are presented in Table 3.

**Table 3.**
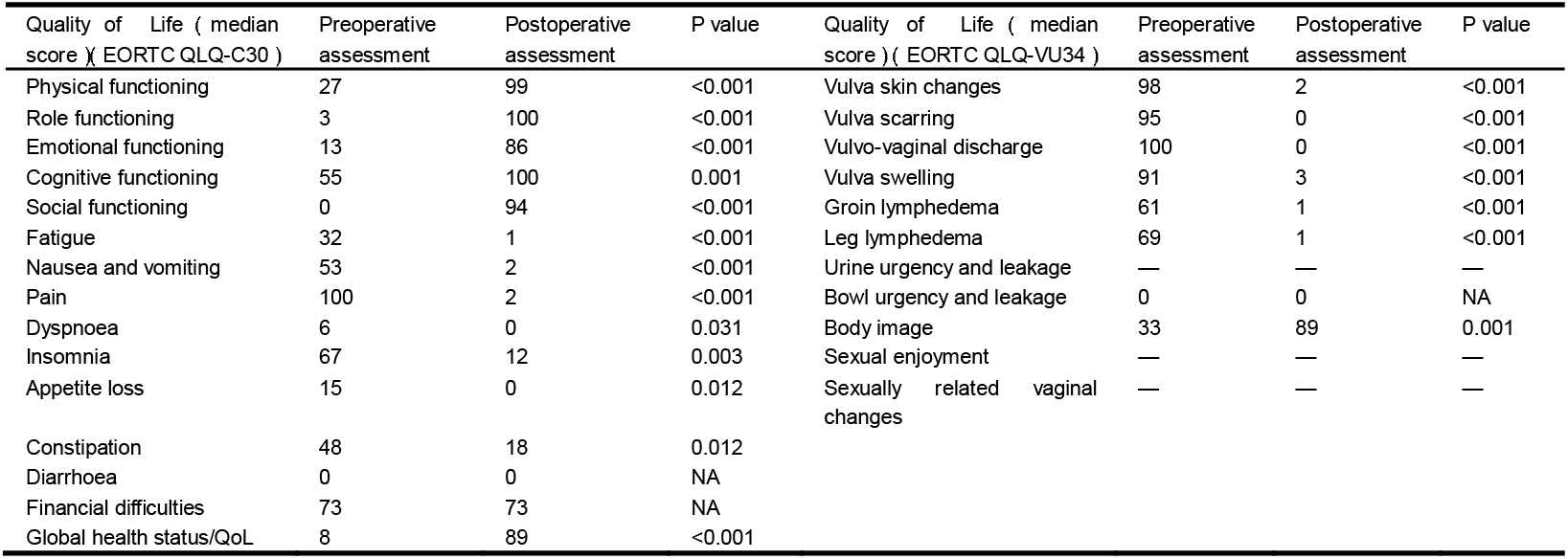
Quality of Life Assessment Before And After Radical Vulvectomy and Rectus Abdominis Flap Reconstruction (EORTC QLQ-C30 V3.0 and EORTC QLQ-VU34)

## Discussion

In this study, we included 11 patients with recurrent vulvar cancer, all of whom received radiation therapy at the time of initial treatment, which is consistent with the use of radiation therapy in the treatment of vulvar cancer. Currently, the main treatment strategies for vulvar cancer include surgical excision and radiotherapy. Surgical excision, the traditional treatment for vulvar cancer, is performed in various ways, and the specific surgical methods need to be determined according to the depth of tumor infiltration and the location of the tumor. Surgical resection can completely remove the tumor tissue and achieve negative margins, but it may result in significant vulvar defects and large perineal scar, which may affect patients’ postoperative quality of life.^1,20,21^

In recent years, with the in-depth research on radiotherapy, several studies have confirmed that radiotherapy has a good effect in the treatment of vulvar cancer, which can significantly reduce the extent of surgical resection or even avoid surgery. ^22-25^Therefore, the application of radiotherapy in the treatment of vulvar cancer has become more and more extensive and has gradually become one of the standard treatment options. In particular, combined radiotherapy and chemotherapy regimens have been shown to provide significant clinical outcomes for patients with vulvar cancer.^26-30^ For example, the GOG 101 trial clinically evaluated radiotherapy in combination with chemotherapy in 46 patients with vulvar squamous cell carcinoma (SCC) with N2/N3 lymph node involvement. The results of the study showed that 38 patients had disease that was resectable following radiotherapy with a cisplatin/5-fluorouracil (5-FU) regimen and subsequently underwent surgery. Local control of lymph node disease was achieved in 36 of 37 patients and control of the primary tumor was achieved in 29 of 38 patients.^31^ However, a large number of studies have found that radiotherapy can cause a variety of complications in the skin in the irradiated area, including vascular damage, decreased cell regeneration, increased inflammatory response, and skin fibrosis, etc. These complications often significantly increase the morbidity and mortality of patients with vulvar cancer. These complications significantly increase the risk of postoperative incisional infection, delayed healing, and non-healing when the patient undergoes reoperation, which can have a significant impact on the patient’s quality of life after surgery. The causes and mechanisms of these complications have been studied to some extent, and radiotherapy techniques have also been significantly improved, but these complications are still a problem that should not be ignored in the treatment process. Therefore, in the surgical treatment of patients with recurrent vulvar cancer after radiotherapy, complications caused by radiotherapy should be emphasized and taken into account in the treatment decision. This requires us not only to focus on local tumor control and survival when making treatment plans, but also to further explore how to optimize the surgical treatment plan to reduce the occurrence of these complications.

In a multicenter case study of 502 patients with vulvar cancer, researchers analyzed recurrence and found that 187 patients (37.3%) experienced recurrence after initial treatment. ^32^Another study of patients with vulvar cancer further revealed the distribution of patients’ time to recurrence, with the majority of recurrences occurring within 2 years of initial treatment, but a subset of patients experienced recurrence after 5 years. ^9,33^These data underscore that vulvar cancer carries a high risk of recurrence after initial treatment and highlight the need for long-term surveillance of patients. There is a lack of clear treatment guidelines for the management of recurrent vulvar cancer, particularly in patients who have received radiotherapy. According to the 2024 edition of the NCCN Guidelines for the Management of Vulvar Cancer, localized vulvectomy and radiation therapy are effective treatments for patients with locally recurrent vulvar cancer who have not been treated with radiation therapy, and radiation therapy alone may also provide some disease-free survival (DFS). ^1,34,35^For patients with locally recurrent vulvar cancer who have received prior radiation therapy, guidelines recommend partial or total radical vulvectomy when conditions permit. This procedure allows for more complete removal of the recurrent tumor and its invading pubic, urethral, vaginal, and rectal tissues to achieve negative margins, but it tends to result in larger perineal tissue defects. In this study, all 11 patients achieved negative margins after radical vulvectomy, but the patients had a median skin defect area of 82.5 cm^2^. This large tissue defect not only increases the risk of wound infection, non-healing, or delayed healing, but also has a significant impact on the psychological and mental health of patients. Therefore, radical vulvectomy is a viable treatment option for patients with recurrent vulvar cancer, especially those with recurrence after radiotherapy, but an individualized postoperative trauma repair plan needs to be developed to protect the patient’s postoperative quality of life.

Due to the unique location of vulvar cancer, defect repair in the perineal area is more challenging than in other areas. Traditionally, surgeons tend to manage postoperative incisions with direct closure, but this method is often difficult to completely close large perineal defects and is associated with the risk of wound dehiscence and scarring. To effectively manage perineal complications after radical vulvectomy, surgeons have proposed a variety of flap reconstruction methods, including the anterior lateral femoral muscle flap, thin femoral muscle flap, gluteus muscle flap, and rectus abdominis muscle flap, each of which has its own advantages and limitations. In this study, we chose a tipped rectus abdominis muscle flap to repair perineal defects after radical vulvectomy. Compared with other types of flaps, the rectus abdominis muscle flap is preferred because of its rich blood flow, easy treatment of the donor area, simple surgical operation, good ductility and healing ability, abundant muscle mass, and more aesthetic appearance. Blood supply to the rectus abdominis flap is primarily dependent on the upper and lower abdominal wall arteries, and this anatomic feature allows one side of the vascular pedicle to be used to support a larger area of the rectus abdominis musculocutaneous flap during flap reconstruction, effectively repairing a larger area of perineal defects. The abdominal wall donor area is relatively easy to retract and suture, allowing direct suturing of the donor area for repair. Although it has been suggested that removal of a rectus abdominis muscle may decrease abdominal wall tension and thus increase the risk of postoperative incisional hernia, in this study, the donor area was repaired by intraoperative use of a patch, and postoperative follow-up showed that none of the 11 patients had donor area complications. In addition, the simplicity of rectus abdominis muscle flap reconstruction and the lack of intraoperative repositioning helped to minimize operative time, with a median operative time of 210 minutes for all patients in this study. The good ductility of the rectus abdominis muscle flap allows it to fill larger areas of perineal defects, and because the rectus abdominis muscle is not in the area of vulvar cancer radiotherapy, it does not suffer from radiologic damage and has a good healing ability, as evidenced by the median postoperative hospital stay of the patients in this study of 24 days. The rectus abdominis muscle flap also has abundant muscle mass, and although the flap may be bulky postoperatively, it can be cosmetically improved after six months due to wasting atrophy of the rectus abdominis muscle. Therefore, the rectus abdominis muscle flap shows its unique advantages in the repair of perineal defects after radical vulvectomy, which not only improves the success rate of the repair, but also helps to improve the patient’s postoperative quality of life, and these advantages provide strong support for the postoperative functional reconstruction of vulvar cancer patients.

In this study, our team performed radical vulvectomy and rectus abdominis muscle flap reconstruction in 11 patients and evaluated the patients’ preoperative and postoperative quality of life. The results showed that patients with recurrent vulvar cancer after radiotherapy had more severe clinical symptoms and very poor quality of life. Their main clinical symptoms included severe perineal pain, perineal ulceration and bleeding, abnormal perineal discharge and odor. In addition, patients’ physical functioning, role functioning, emotional functioning, social functioning, and overall health scores were all at low levels, which clearly showed the poor quality of life of patients with recurrent vulvar cancer after radiotherapy. Therefore, it is essential to provide a treatment program that can improve the quality of life of patients with recurrent vulvar cancer after radiotherapy. In this study, 11 patients were followed for a long period of time and their postoperative quality of life was reassessed. At the last follow-up, the median follow-up time in this study was 13 months, and 10 of the 11 patients remained disease-free and survived, while one patient developed localized intra-pelvic recurrence, indicating that the treatment regimen of radical vulvectomy and rectus abdominis muscle flap reconstruction had relatively good therapeutic efficacy. Meanwhile, the postoperative quality of life scores showed that a series of quality of life scores, including perineal pain, perineal ulcer, perineal odor, and general health, underwent significant improvement, which indicated that the patients’ postoperative quality of life was greatly improved, and further illustrated the good effect of radical vulvectomy and rectus abdominis muscle flap reconstruction in the treatment of recurrent vulvar cancer after radiotherapy.

In this study, we demonstrated the feasibility and good results of radical vulvectomy and rectus abdominis muscle flap reconstruction for the treatment of recurrent vulvar cancer after radiotherapy through a relevant follow-up study. However, there are several limitations of this study; as a retrospective study, we have the disadvantages of small sample size, short follow-up time, and lack of randomized controlled trials, which may limit our direct assessment of the superiority of treatment. In the future, we will continue to follow the enrolled patients for a long period of time to build a comprehensive follow-up database, which will provide richer data support for further research on vulvar cancer treatment. Through continuous data collection and analysis, we expect to be able to more comprehensively evaluate the effectiveness of radical vulvectomy and rectus abdominis flap treatment, and provide more scientific basis for the treatment of vulvar cancer patients.

## CONCLUSIONS

Radical vulvectomy and functional reconstruction of rectus abdominis muscle flap for the treatment of recurrent vulvar cancer after radiotherapy is an effective and reliable surgical plan, which has the advantages of shorter operation time, less bleeding, negative margins, and faster postoperative recovery time. At the same time, this treatment program can significantly improve the clinical symptoms of patients and thus significantly improve the quality of life of patients. However, we do not recommend that surgeons without relevant experience perform this procedure independently, as it is difficult to operate and requires not only specialized episiotomy techniques but also relevant plastic surgery skills. Therefore, we recommend that surgeons and plastic surgeons work together to perform this procedure when conditions permit.

## Data Availability

All data produced in the present study are available upon reasonable request to the authors

